# From Black Box to Discovery Engine: A Geometric and Topological Framework for Interpreting Graph Neural Networks in Critical Care

**DOI:** 10.1101/2025.08.06.25333083

**Authors:** Sohyeon Jeon

## Abstract

Effective clinical decision-making in critical care depends on interpreting complex, high-dimensional patient data. However, many advanced AI models function as uninterpretable “black boxes,” limiting their trustworthiness and scientific value. This study introduces NETWISE, a geometric and topological framework designed not for mere prediction, but for building a validated patient similarity network to serve as a scientific discovery engine. We demonstrate the framework’s utility using acute cholangitis, a life-threatening emergency, as a case study. Analyzing 1,372 patients from the MIMIC-IV database, NETWISE constructs a heterogeneous graph to learn a robust patient manifold. The resulting model achieved high predictive accuracy (AUC-ROC of 82.5%) while offering multiple layers of interpretability. The framework’s potential for discovery was demonstrated through two complementary analyses: SHAP identified clinically coherent predictors, while discrete clustering autonomously generated 13 data-driven patient subgroups, presenting them as testable hypotheses for future clinical investigation. Crucially, topological analysis confirmed the network’s immense structural complexity (β1 = 63,187). This study’s primary contribution is a transparent, validated, and network-based methodological framework capable of generating new, data-driven clinical hypotheses from complex EHR data.

## 1. Introduction

Critical care medicine operates at the intersection of high-stakes decision-making and profound data complexity [58,59]. Clinicians must navigate a deluge of high-frequency physiological data, laboratory results, and clinical notes to manage patients with rapidly evolving, life-threatening conditions [60]. Acute cholangitis serves as a compelling archetype of this challenge: a life-threatening biliary infection that can rapidly escalate to sepsis and multi-organ failure, demanding swift and precise decision-making in high-stakes environments like the emergency department (ED) and intensive-care unit (ICU) [1–3, 11,12]. The most severe cases (Grade III) carry a 30-day mortality rate as high as 36%, making accurate triage for timely intervention a matter of survival [4–7,13–15]. However, clinicians face a fundamental challenge: the profound heterogeneity of patient trajectories. While foundational and invaluable for standardized risk assessment, conventional prognostic tools—from simple nomograms to standard critical-care scores like SOFA and APACHE II—are not inherently designed to model the relational context in which clinical events unfold [62–64]. By processing patients as independent entities, these methods are limited in their ability to capture the complex web of similarities between patient trajectories or the high-order interactions among evolving clinical states [8, 32–36]. This gap represents a critical opportunity for a new class of analytical methods that can complement existing tools by embracing this real-world complexity.

To address this, Graph Neural Networks (GNNs) have emerged as a powerful paradigm. Electronic health records are inherently relational—linking patients, diagnoses, treatments and outcomes—yet such structure is poorly handled by tabular algorithms [16–18]. By representing electronic health records (EHRs) as heterogeneous graphs, GNNs can model the intricate web of relationships between patients, diagnoses, and treatments, demonstrating state-of-the-art predictive performance across numerous clinical tasks [21–26, 37–44]. Despite their predictive power, their adoption in routine clinical care is hindered by a critical barrier: their ‘black-box’ nature. This lack of transparency creates a profound gap between technological innovation and practical application. For clinicians to trust and act upon an AI’s recommendation, especially in life-or-death situations, they need more than an accurate prediction; they need to understand the reasoning behind it. This gap is the central obstacle preventing advanced AI from realizing its full translational potential.

Existing eXplainable AI (XAI) methods, such as SHAP (SHapley Additive exPlanations), have attempted to bridge this gap by identifying which input features are most important for a given prediction [61]. While valuable, this approach offers only a partial view. It answers ‘what’ variables the model is looking at but fails to illuminate the high-level knowledge structure the model has learned from those variables. It cannot tell us how the model organizes patient relationships or conceptualizes disease progression. This leads us to a more fundamental question, moving beyond conventional XAI: “What is the high-level clinical knowledge structure that a GNN autonomously learns from complex medical data? Is this learned knowledge simply a collection of discrete patient clusters, or does it possess a more intricate, a continuous manifold with non-trivial topology?” All modern AI ultimately ingests tensors, but the route from raw data to those tensors is architecture dependent. Language models tokenize notes into embedding matrices; CNNs rasterize waveforms into image-like grids. In our approach, the same EHR is translated into a heterogeneous graph whose nodes (e.g. patients, labs, diagnoses) and edges (e.g. clinical events, similarity links) preserve the relational fabric of critical care. This representation allows learning from both individual physiology and population-level structure yet also deepens the interpretability challenge.

The central challenge in clinical AI, therefore, is not merely to build more accurate predictive models, but to develop validated frameworks that transform complex patient data into an interpretable manifold for scientific discovery [30,31]. To address this, this study introduces NETWISE (Network-Enhanced Treatment and Warning Intelligence System for Emergencies), a modular, disease-agnostic framework that moves beyond a single prediction model. It is crucial to define the scope of this paper: our primary objective is to introduce and validate the NETWISE framework itself—its architecture, its computational validity, and its capacity to generate interpretable structures. The resulting outputs, such as patient clusters or identified hub nodes, are presented not as definitive clinical discoveries, but as concrete examples of data-driven hypotheses that this engine can produce. The essential subsequent step of clinical validation is intentionally left for future interdisciplinary work. Using acute cholangitis cases from the MIMIC-IV database as a proof-of-concept, we demonstrate a principled approach to (i) construct a topologically complex patient similarity network, (ii) train a heterogeneous GNN that learns clinically coherent patterns, and (iii) validate the learned structure using a multi-scale geometric-topological interpreter. The modular design of NETWISE ensures that its core graph engine is adaptable to a broad array of clinical scenarios, facilitating rigorous, reproducible research and advancing the translational potential of network-based precision medicine.

## 2. Methods

### 2.1. Study Design and Dataset

This retrospective cohort study developed and validated NETWISE using the publicly available, de-identified Medical Information Mart for Intensive Care IV (MIMIC-IV) database [27]. Adult patients (18–80 years) with acute cholangitis and related biliary complications were identified using a comprehensive set of 30 ICD-9 and 78 ICD-10 codes, based on the diagnostic criteria of the Tokyo Guidelines 2018 [13–15, 44]. These codes encompassed not only primary acute cholangitis but also related conditions including cholelithiasis with cholangitis, biliary obstruction, and associated sepsis.

Inclusion criteria were strictly defined to ensure a high-quality analytical cohort. A hospital stay of at least 24 hours and a minimum of one ICU admission were required. To maintain consistency and avoid confounding effects from multiple admissions, only the first ICU stay per patient was retained for analysis. Patients were excluded if any essential demographic or admission data (e.g., patient identifiers, age, admission times) were missing, ensuring a complete foundational record. From an initial screening of 8,306 admissions, 1,372 unique patients constituted the final analytical cohort.

To prevent data leakage, strict temporal separation was enforced: predictors were limited to information available during the first 24 h of ICU stay, whereas the outcome—72-hour clinical deterioration— was defined as a composite endpoint including the occurrence of one or more of the following events within the subsequent 72-hour window: initiation of vasopressor therapy, initiation of mechanical ventilation, or the onset of new oliguria.

### 2.2. Data Preprocessing and Feature Engineering

The study constructed two distinct feature sets for different purposes: a comprehensive set for the predictive model and a curated set for constructing the patient similarity network. All feature engineering was conducted on data from the first 24 hours of ICU admission.

For the primary prediction task, a set of 16 clinical features was engineered for each ICU Stay node based on established prognostic frameworks from recent machine learning studies in acute cholangitis [20,29,33]. This selection was based on established prognostic frameworks from recent studies in acute cholangitis. Raw temporal data, including 21 laboratory parameters and 5 vital signs, were processed using temporal aggregation functions (mean, maximum, minimum). While established practices for handling missing data in EHRs include univariate methods like median substitution for laboratory parameters and forward-fill imputation for temporal vital signs [9,10,19,28], these approaches have limitations in capturing complex clinical relationships. Therefore, to handle missing data, we employed IterativeImputer from scikit-learn, which models each feature with missing values as a function of others, thereby preserving inter-variable correlations. The final 16-variable feature set for prediction comprised:

WBC_max, hemoglobin_min, MCV_max, platelet_min, AST_max, ALT_max, alkaline_phosphatase_max, bilirubin_max, direct_bilirubin_max, albumin_min, RDW_mean, RDW_max, PTT_mean, MCHC_mean, chloride_mean, and phosphate_max.

To construct a structurally meaningful patient similarity network, a separate, curated set of 14 variables was assembled. This set was designed to capture a holistic patient profile by combining information from three distinct domains: demographic, social, and clinical. The variables included are detailed below and were used to create a unified feature vector for each patient for similarity calculation.

### 2.3. NETWISE System Architecture

The NETWISE system architecture is designed around a two-stage process that separates persistent data storage from the computational model. First, comprehensive patient data from MIMIC-IV is modeled and stored in a Neo4j graph database, which serves as a robust and quaryable backend. Second, our data processing pipeline queries this database to dynamically construct a task-specific, in-memory heterogeneous graph G = (V, E, τ, *φ*) for direct consumption by the GNN model.

The framework incorporates eight distinct node types A = {patient, admission, icu_stay, lab_event, lab_item, chart_event, item, diagnosis} with |V| = 140,321 total nodes. Node types include: (i) Patient nodes (n=1,372) containing demographic information, (ii) Admission nodes with hospital stay metadata (n=1,372), (iii) ICU Stay nodes serving as primary prediction targets with aggregated clinical features (n=1,372), (iv) Lab Event nodes with individual laboratory test results and temporal information (n=46,494), (v) Lab Item nodes containing laboratory test categories and reference ranges (n=21), (vi) Chart Event nodes with vital signs and monitoring data (n=87,901), (vii) Item nodes with chart event categories and measurement units (n=5), and (viii) Diagnosis nodes with ICD-coded diagnoses (n=1,784).

The graph contains four primary edge categories: (1) Medical Workflow Edges capturing patient care trajectories through patient → admission → icu_stay paths and diagnostic procedures via admission → {lab_event, diagnosis} connections, (2) Definition Edges linking lab_event ↔ lab_item and chart_event ↔ item for test type associations, (3) Clinical Similarity Network with |E_similarity| = 135,382 patient-to-patient connections based on cosine similarity across standardized feature space, representing ≈51.3% of the total graph connectivity and enabling comprehensive collective pattern learning, and (4) Enhanced Medical Relationships: These edges capture fine-grained clinical events. For instance, an edge connects the lab_item ‘Creatinine’ with the chart_item ‘Urine Output’ to represent their physiological relationship. This includes temporal associations linking an icu_stay to the specific lab_events and chart_events (e.g., vital sign measurements) that occurred during that stay. It also incorporates connections between clinically related lab_item and item nodes. All relationships include bidirectional edges to capture symmetric dependencies.

#### Formal Graph Definition

The heterogeneous graph is formally defined as ***G*** = (***V, E, X, τ, φ***) where:

- *V* = *V*_*patient*_ ∪ *V*_*admission*_ ∪ *V*_*icustay*_ ∪ *V*_*labevent*_ ∪ *V*_*labitem*_ ∪ *V*_*chartevent*_∪ *V*_*item*_ ∪ *V*_*diagnosis*_ represents the union of eight node type sets with |V| = 140,321 total nodes
- *E* = *E*_*workflow*_∪ *E*_definition_ ∪ *E*_similarity_ ∪ *E*_enhanced_ represents the union of four edge categories with |E| = 263,837 total edges
- *X*: *V* → ℝ^*d*^ denotes the node feature mapping function (d varies by node type)
- τ: *V* → *A* defines the node type mapping function to A = {Patient, Admission, ICUStay, Labevent, Labitem, Chartevent, Item, Diagnosis}
- *φ*: *E* → *R* defines the edge type mapping function to relation types *R* including medical workflows, clinical similarities, and enhanced relationships

This graph structure is enriched by a patient similarity network (PSN), connecting patientnodes based on their holistic clinical profiles. The similarity was computed not on the predictive features alone, but on the dedicated 14-variable vector described in Section 2.2. This vector integrates information from three weighted domains to ensure a balanced representation of patient characteristics, as detailed in Table I.

**Table I.** Feature Domains FOR Patient Similarity Calculation.

| Domain | Weight | Variables |
| --- | --- | --- |
| Demographic | 0.1 | age, gender |
| Social & Administrative | 0.2 | race, marital_status, insurance, admission type |
| Clinical | 0.7 | hospital_stay_hours, bilirubin_max, alt_max, crp_max, wbc_max, fever_high (derived), elderly (derived), severity_score (derived) |

Each feature subset was independently standardized (Z-score normalization). The final feature vector xi for each patient i was constructed by concatenating these standardized subsets, with the contribution of each subset scaled by its respective weight. Similarity between two patients i and j was then calculated using cosine similarity: similarity (i, j) = (xi · xj) / (||xi|| ||xj||). An edge was created between two patients i and j if their cosine similarity exceeded a threshold of 0.85, and if each patient was within the other’s top 5 nearest neighbors (k=5), thus forming a robust backbone for the PSN.

### 2.4. Model Training and Validation

We performed a patient-level stratified split to prevent information leakage, allocating 70% of the cohort (n=960) for training, 10% (n=137) for validation, and 20% (n=275) for testing, based on the final analytical cohort of 1,372 patients. All splits preserved the primary endpoint —72-hour clinical deterioration—to maintain class balance.

Hyperparameters were selected through five-fold stratified cross-validation on the development set (training and validation), targeting sensitivity ≥80% and specificity ≥70%. The final model is a two-layer graph attention network (GAT) with 128 channels in the first layer and 64 in the second, utilizing eight attention heads per layer. The model was trained using a multi-task learning framework on five objectives: node-level deterioration prediction, link prediction, community detection, and spectral-topological regularization. To address class imbalance in the primary prediction task, a focal loss function was employed (α = 0.3, γ = 2.5). The output from the final GAT layer is then passed through a linear projection layer to generate the final 512-dimensional node embeddings used for downstream analysis.

The model incorporated several regularization techniques to prevent overfitting, including a dropout rate of 0.4, weight decay of 0.02, and gradient-norm clipping at 1.0. The learning rate was dynamically managed by a Reduce-LR-on-Plateau scheduler, which reduced the rate by a factor of 0.7 after 15 epochs without improvement. Training was halted at epoch 330 by the early stopping mechanism (patience = 30) after the validation AUC failed to show further improvement. During training, task weights were adaptively adjusted; the final weights for the primary node classification, spectral loss, topological loss, link prediction, and community detection tasks were 31.67, 0.05, 0.14, 0.20, and 0.10, respectively. Model performance was assessed on the test set using AUC-ROC, sensitivity, specificity, accuracy, positive predictive value (PPV), negative predictive value (NPV), and F1-score. Results are reported for both the default threshold (0.50) and the Youden-optimized threshold to provide a comprehensive view of the model’s performance at different clinical operating points.

### 2.5. Explainability Analysis

To assess model interpretability, we employed SHAP to estimate the contribution of each input variable to the model’s predictions. Given the computational demands of explaining a graph-based model, we utilized the KernelExplainermethod from the shap library. The explainer was constructed using a background dataset of 50 samples randomly selected from the training set. Global feature importance was then estimated by calculating SHAP values for 20 representative cases from the test set, ensuring stable and robust estimates for the summary analysis. This analysis focuses on explaining the primary predictive task of the model.

### 2.6. Validation and Interpretation of the Learned Latent Space

Beyond predictive performance, we conducted a multi-scale post-hoc analysis on the final node embeddings to validate the quality of the representations learned by the GNN. This two-part validation was designed to assess whether the model successfully organized the complex patient data into a mathematically robust and clinically interpretable manifold, thereby serving as a validated engine for scientific discovery.

To determine if the GNN could autonomously identify structurally distinct patient subgroups based on the learned embeddings, we performed unsupervised clustering on the learned embeddings. First, we extracted the final 512-dimensional embeddings of all ICU Stay nodes. The high-dimensional embeddings were then projected into a lower-dimensional space using Uniform Manifold Approximation and Projection (UMAP). Following this, Density-Based Spatial Clustering of Applications with Noise (DBSCAN) was applied to the reduced-dimensional data to identify distinct patient clusters. The clinical characteristics of any resulting data-driven clusters would then be explored by comparing the distributions of input features by comparing the distributions of all 16 input features across the clusters using appropriate statistical tests (e.g., Kruskal-Wallis).

To validate the structural integrity of the representations learned from the full heterogeneous graph, we constructed and analyzed a specific Patient Similarity Network (PSN). This PSN was derived from the final patient node embeddings generated by the GNN and serves as a human-interpretable projection of the high-dimensional latent space. This validation was performed using two complementary approaches:

First, to assess the topological complexity of the PSN, we quantified the number of independent cycles by calculating its first Betti number (*β*1). This computation was performed using specialized topological data analysis (TDA) libraries, primarily torch-topological, to efficiently handle the large-scale graph structure. This metric measures the presence of non-trivial cyclical relationships between patients that cannot be captured by simple hierarchical structures. Second, to confirm the statistical stability of this complex topology, we employed a bootstrap validation procedure. This involved repeatedly creating subgraphs by sampling 90% of the patient nodes without replacement for 1,000 iterations. For each subgraph, the first Betti number was recalculated. The mean and 95% confidence interval of *β*1 were then derived from this distribution of 1,000 values to ensure that the observed topological complexity was a robust feature of the learned manifold and not a random artifact.

### 2.7. Machine Learning Baseline Models

To establish comprehensive performance benchmarks and validate the superiority of the graph-based approach, eight traditional machine learning models were systematically implemented and evaluated using identical data preprocessing and temporal separation protocols.

The baseline models included Logistic Regression, a Support Vector Machine with RBF kernel, Naive Bayes, a Multi-Layer Perceptron (128-64 hidden layers), Random Forest, and two implementations of Gradient Boosting: XGBoost (v3.0.2) and LightGBM (v4.6.0). All models were configured with random seed 42 for reproducibility and commonly used default hyperparameters to ensure fair comparison. Traditional ML models utilized the same features as the GNN model, extracted from ICU stay node features. The tabular data underwent systematic preprocessing including median imputation for missing values and standardization using StandardScaler. Class imbalance was addressed through balanced class weights for applicable models when the training positive ratio fell outside the 30-70% range.

All models were trained using identical patient-level data splits (70% training, 10% validation, 20% testing) to ensure no data leakage between training and evaluation sets. Model performance was assessed using the same comprehensive metrics as the GNN model, with optimal classification thresholds determined using the Youden Index for each model independently. Statistical significance was assessed using DeLong’s test for AUC comparison and McNemar’s test for paired predictions. NETWISE was compared against eight traditional machine learning baselines trained on identical datasets with the same temporal separation protocol.

### 2.8. Computational Environment

All data processing, model training, and analyses were conducted on a MacBook Pro equipped with an Apple M4 Pro chip (14-core CPU), 48 GB of unified memory, and running macOS. The computational environment was managed using Conda (version 24.5.0). Key software packages included Python (3.11.13), PyTorch (2.4.0), PyTorch Geometric (2.6.1), and Scikit-learn (1.6.1). For specific analyses, we utilized shap (v0.48.0) for feature interpretability and torch-topological for the topological validation.

## 3. Results

### 3.1. Patient Characteristics and Study Population

The final analytical cohort comprised 1,372 patients meeting all inclusion and exclusion criteria from the MIMIC-IV database. The patient selection process successfully identified a representative acute cholangitis population with a mean age of 61.0 ± 12.9 years and 58.3% male patients. All patients had an ICU admission with a hospital length of stay ≥24 hours, ensuring adequate clinical data capture for the 24-hour prediction window. For model evaluation, the test set (n=275) was well-balanced, with the primary outcome of 72-hour clinical deterioration occurring in 138 patients (50.2%). Patient characteristics were consistently distributed across the training, validation, and test sets, with no significant differences observed (p>0.05 for all comparisons).

### 3.2. Predictive Performance and Patient Manifold Validation

NETWISE demonstrated strong predictive performance, which was evaluated at two distinct operating points. At the default threshold (0.50), the system achieved a sensitivity of 73.9%and a specificity of 71.5%. The optimal threshold determined by the Youden Index (0.524) provided a balanced performance profile with a sensitivity of 71.7% and a specificity of 73.7%. Both scenarios are underpinned by a robust and consistent AUC-ROC of 82.5%. Detailed performance metrics are presented in Table II. The training was halted by the early stopping mechanism at epoch 330 after the validation AUC failed to improve for 30 consecutive epochs, indicating the model reached optimal performance without overfitting.

**TABLE II.** NETWISE Performance AT Different Clinical Threshold.

| Metric | High-Sensitivity<br>(Threshold: 0.5) | Balanced (Youden Index,<br>Threshold: 0.524) |
| --- | --- | --- |
| AUC-ROC | 82.5% | 82.5% |
| Sensitivity | 73.9% | 71.7% |
| Specificity | 71.5% | 73.7% |
| Accuracy | 72.7% | 72.7% |
| F1-Score | 73.1% | 72.5% |
| PPV | 72.3% | 73.3% |
| NPV | 73.1% | 72.1% |

To validate that the GNN learned a clinically meaningful and structurally robust representation, we analyzed the final Patient Similarity Network (PSN) constructed from the learned patientnode embeddings. The analysis confirmed the emergence of a complex, non-trivial structure, moving beyond the ad-hoc nature of traditional network construction methods. The resulting network connects 1,372 patients with 64,558 similarity edges, forming a single, fully connected component. The network exhibits a density of 6.86% and a high average clustering coefficient of 0.593, indicating that patients are not isolated but are organized into dense, tight-knit local communities, akin to a complex social network.

Crucially, the network’s topological complexity was quantified by its first Betti number (*β*1), which was 63,187. To confirm the statistical stability of this finding, the bootstrap validation described in the methods yielded a mean *β*1 of 51,014 with a tight 95% confidence interval of [49,321 – 52,577]. The narrowness of this interval, derived from 1,000 resamples, provides strong evidence that this immense topological complexity is a robust and stable feature of the learned manifold, and not a random artifact. This high value reveals the existence of tens of thousands of independent cyclical relationships between patients. It provides strong evidence that the GNN captured more than simple pairwise similarities, learning a rich manifold of high-order, non-hierarchical patient interdependencies that would be invisible to traditional methods.

This complex structure gives rise to structurally significant topological features, such as hub nodes. For example, patient 817 emerged as the top hub, connecting directly to 327 other patients and acting as a central anchor in the network’s densest region (Fig. 1). To illustrate this, a post-hoc analysis of the top hub, Patient 817, provides a concrete example of hypothesis generation. This patient presented with a risk score of 0.923, placing them in the highest risk tier. More strikingly, their maximum C-reactive protein (CRP) level was 3364.0 mg/L, a value more than ten times the cohort average (310.8 mg/L). This profile suggests that Patient 817 could function as a computational prototype for a potential hyper-inflammatory subgroup, generating a clear, testable hypothesis. The model autonomously identified this patient as a structural anchor, thus generating a data-driven hypothesis about the central role of severe inflammation in a major patient subgroup, which merits further clinical validation. Complementing this analysis of individual hubs and continuous cycles, we next examined the model’s ability to identify discrete patient subgroups. To fulfill the discrete analysis axis of our framework, we applied a UMAP and DBSCAN clustering pipeline to the learned patient embeddings to examine the model’s ability to identify discrete patient subgroups.

**Fig. 1.**
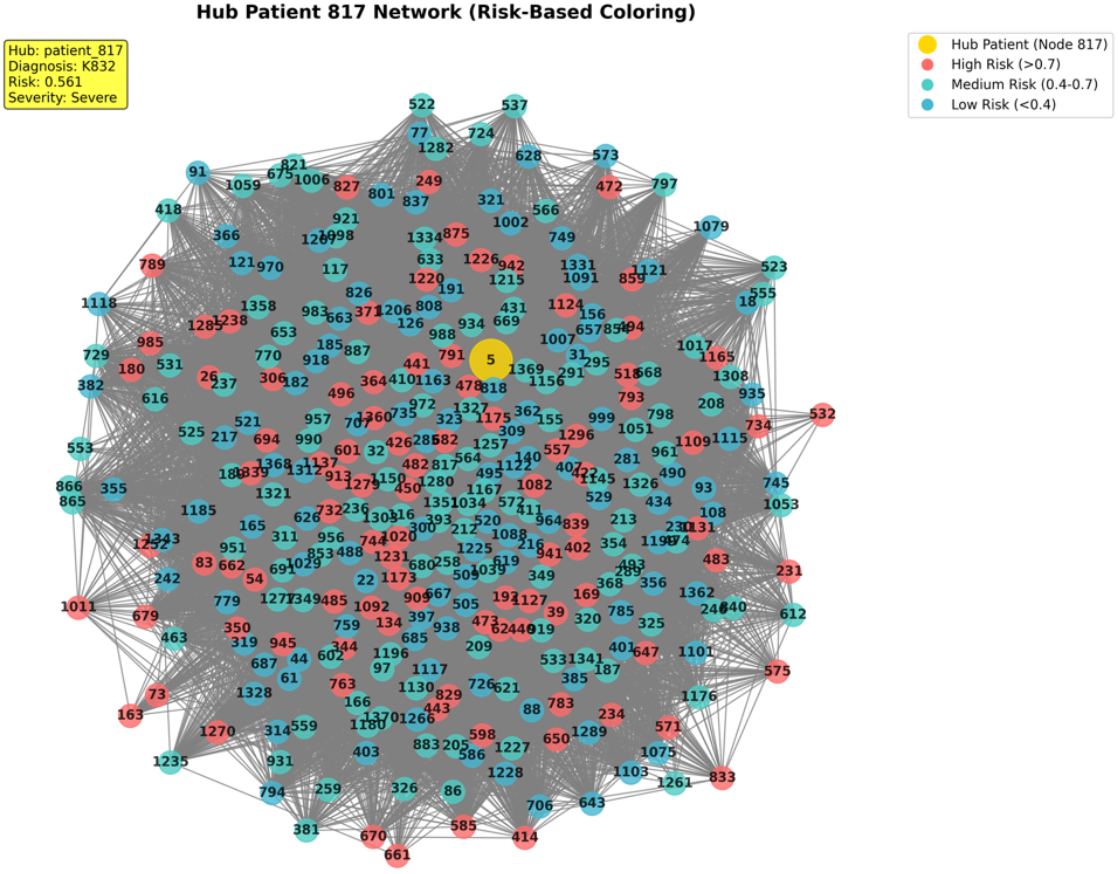
Visualization of the Top Hub Node and Its Local Neighborhood. The network’s most central node, Patient 817 (highlighted in yellow), and its 327 direct neighbors. The density of connections illustrates the formation of a tight-knit community, with the hub acting as a potential computational prototype anchoring the cluster.

The analysis revealed 13 distinct patient clusters from the cohort of 1,372 patients, as visualized in Fig. 2. These data-driven subgroups emerged without any prior clinical labels, demonstrating the model’s ability to learn and segment the underlying structure of patient heterogeneity. The existence of these well-separated clusters provides a discrete structural map to complement the network’s continuous topological features. This validates that the learned patient manifold is not a uniform, undifferentiated space, but is organized into specific, identifiable sub-regions. Interestingly, a post-hoc analysis comparing the input features across these 13 clusters did not reveal statistically significant differences using conventional univariate tests. This suggests that the GNN identified subgroups based on subtle, high-order interactions between variables, creating subgroups that are meaningfully distinct in the high-dimensional latent space but not easily separable by simple, individual clinical metrics.

**Fig. 2.**
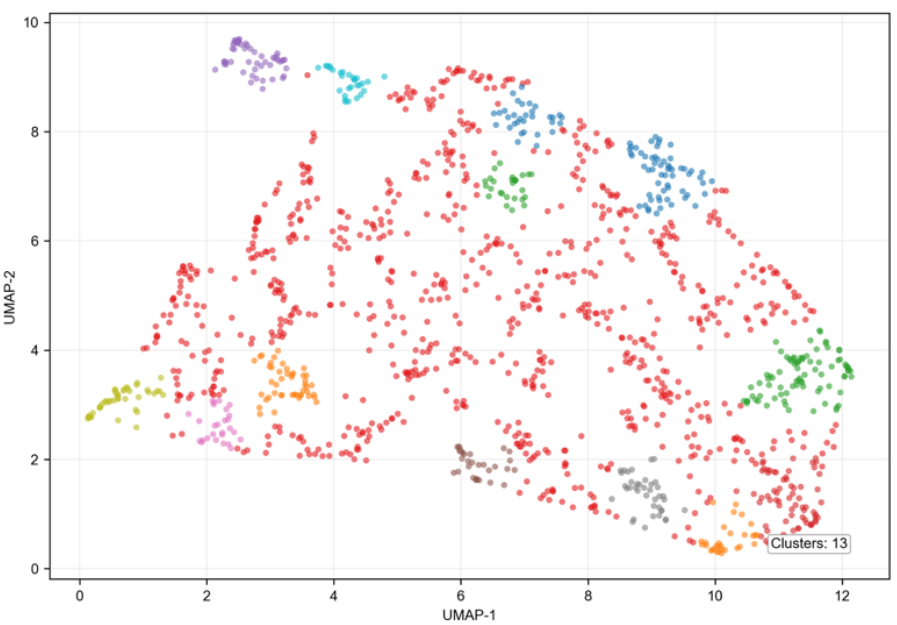
Data-Driven Cluster Discovery in the Patient Latent Space using UMAP. The algorithm identified 13 distinct clusters (colored), representing automatically discovered data-driven subgroups from the 1,372 patient cohort. The remaining points (in red) are outliers not assigned to any specific cluster, potentially representing highly atypical patient profiles that do not conform to the dominant subgroups.

### 3.3. Comparative Analysis: NETWISE versus Traditional Machine Learning

To validate the superiority of the graph-based approach, NETWISE was benchmarked against eight traditional machine learning models trained on the same features and data splits. The comprehensive performance comparison is presented in both Table III and Fig 3.

**Table III.** Comparative Performance OF NETWISE VS Traditional Machine Learning Models.

| Model | AUC | Accuracy | F1-Score | Overfitting Gap<br>(Train AUC – Test AUC) |
| --- | --- | --- | --- | --- |
| NETWISE* | 82.5% | 72.7% | 72.5% | Controlled by early stopping |
| Gradient Boosting | 74.4% | 69.8% | 70.0% | 21.9% |
| XGBoost | 72.3% | 68.0% | 67.2% | 27.7% |
| Random Forest | 72.1% | 67.2% | 67.2% | 27.9% |
| LightGBM | 71.1% | 66.4% | 66.4% | 28.9% |
| SVM (RBF) | 70.9% | 63.4% | 63.4% | 13.2% |
| Logistic Regression | 70.1% | 65.2% | 65.2% | 4.9% |
| Neural Network<br>(MLP) | 68.9% | 63.5% | 63.5% | 5.8% |
| Naïve Bayes | 68.7% | 56.0% | 36.0% | 0.8% |

\*NETWISE performance reported at the Youden Index-optimized threshold. All baseline model metrics are from the test set.

**Fig. 3.**
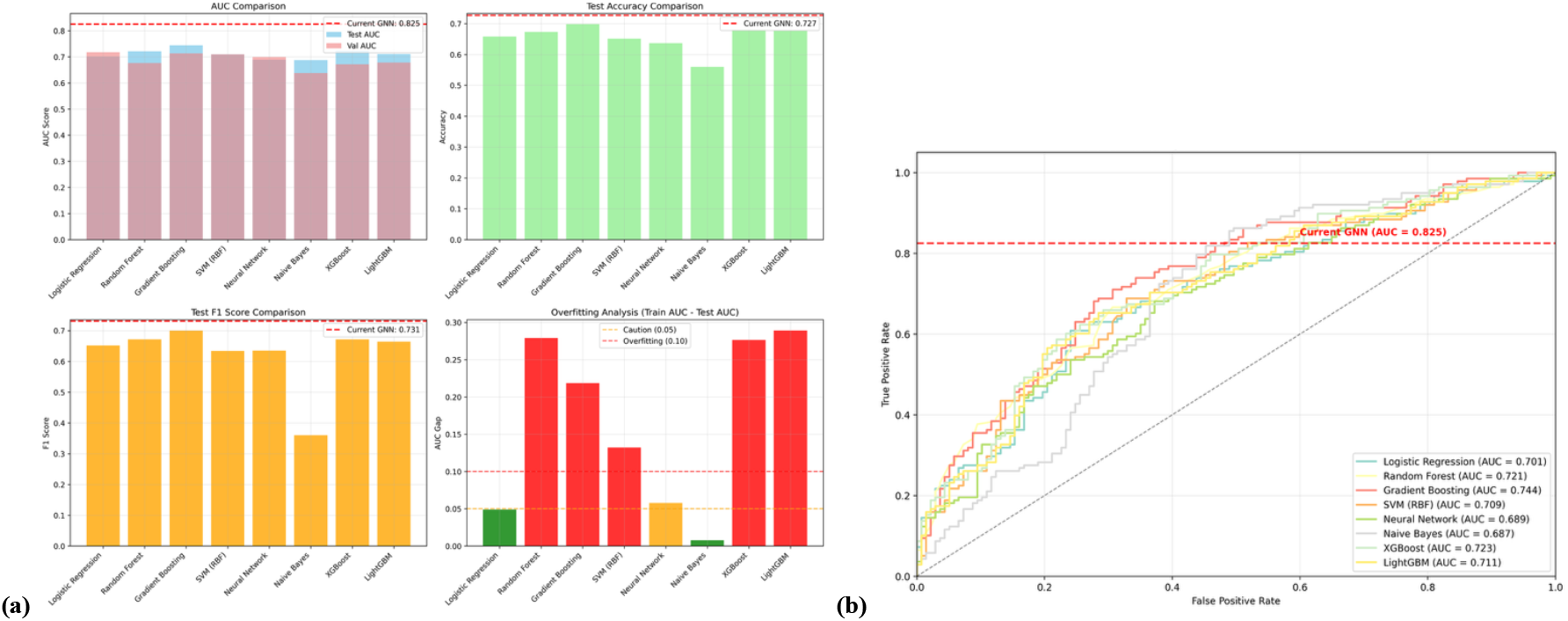
Performance Comparison of NETWISE against Traditional Machine Learning Baselines. **(a)** A comparative analysis across four key metrics: Area Under the Curve (AUC), Accuracy, F1-Score, and an overfitting analysis (Train AUC - Test AUC). The performance of NETWISE is indicated by the red dashed line in each subplot. NETWISE achieved a higher AUC and F1-Score than all baseline models, while also avoiding the significant overfitting observed in high-performing ensemble models like Gradient Boosting and XGBoost. **(b)** Receiver Operating Characteristic (ROC) curves for all evaluated models. NETWISE (AUC = 0.825) exhibits the largest area under the curve, demonstrating its superior discriminative ability in predicting clinical deterioration compared to all traditional models.

Among the traditional models, Gradient Boosting achieved the highest performance with an AUC of 0.744, an accuracy of 69.8%, and an F1-score of 0.700. Other tree-based ensemble models like XGBoost (AUC 0.723) and Random Forest (AUC 0.721) also performed competitively. However, these models exhibited significant overfitting, with training AUCs approaching 1.0 while test AUCs remained much lower, as noted in the experiment log. In contrast, simpler models like Logistic Regression (AUC 0.701) showed better generalization with a minimal overfitting gap.

NETWISE demonstrated markedly superior performance compared to all traditional baselines. Against the best-performing baseline (Gradient Boosting), NETWISE achieved an 8.1 percentage point improvement in AUC (82.5% vs. 74.4%). This substantial gain underscores the value of leveraging the graph’s complex relational structure, a capability that traditional tabular models lack. The robust performance of NETWISE, achieved without the significant overfitting seen in the top-performing baselines, highlights its strong generalization capability and clinical potential.

### 3.4. Feature Importance and Clinical Interpretability

To understand which clinical variables were most influential, we conducted a SHAP analysis, implemented with the shap library (v0.48.0). Mean absolute SHAP values calculated across the test set were used to rank global feature importance, as shown in Fig 4. The five most important features were: (i) wbc_max (importance score: 0.1170), (ii) ast_max(0.0886), (iii) alkaline_phosphatase_max (0.0768), (iv) rdw_mean (0.0746), and (v) albumin_min(0.0726). These findings strongly align with the established pathophysiology of acute cholangitis. Elevated WBC count (wbc_max) is a primary marker for infection and systemic inflammation. High levels of AST (ast_max) and alkaline phosphatase (alkaline_phosphatase_max) indicate cholestatic liver injury, a hallmark of the disease. Elevated red cell distribution width (rdw_mean) reflects systemic inflammatory response, and low albumin (albumin_min) signifies deteriorating liver synthetic function and nutritional status. Individual case analysis further demonstrated that the model’s predictions are driven by these clinically relevant factors, enabling clinicians to understand the specific drivers contributing to each patient’s risk assessment.

**Fig. 4.**
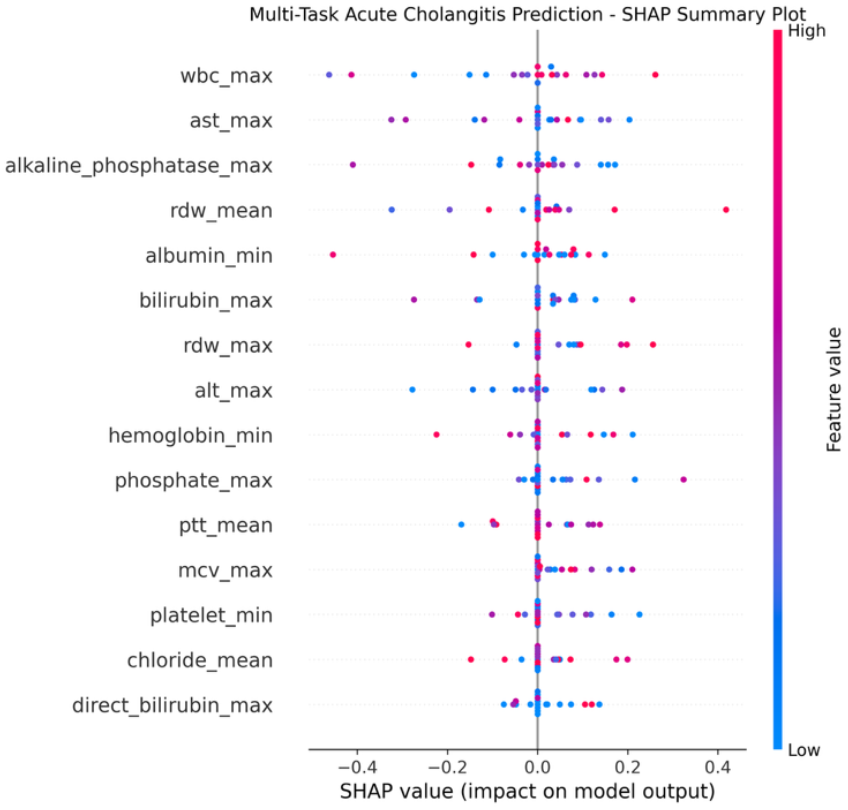
SHAP Analysis of Key Clinical Predictors. The plot summarizes the most influential features for predicting 72-hour clinical deterioration. Features are ranked by their overall impact. For each feature, a single point represents a patient, with its position on the x-axis showing the impact on model prediction (positive values increase risk) and its color indicating the feature value (red for high, blue for low). This reveals, for instance, that high WBC counts (top feature, in red) consistently drive the model to predict a higher risk of deterioration.

## 4. Discussion

In this study, we introduced and validated NETWISE, a framework designed to address a fundamental challenge in clinical AI: moving beyond opaque prediction models to create validated, interpretable frameworks that serve as engines for scientific discovery. Our results confirm that by embracing the inherent relational structure of patient data, we can achieve not only superior predictive performance but also uncover structurally complex topological structures that are invisible to traditional methods.

A primary strength of the NETWISE architecture is its inherent modularity and scalability. While this study focused on acute cholangitis as a proof-of-concept, the framework is designed to be disease-agnostic. The decoupled two-stage architecture allows for remarkable flexibility; by simply modifying the initial patient cohort query in Neo4j (e.g., by selecting a different ICD code) and adjusting the feature selection configuration, this entire discovery pipeline can be readily applied to other critical care conditions such as sepsis, acute respiratory distress syndrome, or cardiogenic shock. This adaptability transforms the framework from a single-purpose tool into a versatile, reusable engine for hypothesis generation across a wide spectrum of diseases.

The principal finding is that NETWISE significantly outperforms eight traditional machine learning baselines, achieving an AUC of 82.5%—a substantial 8.1 percentage point improvement over the best-performing baseline. This performance gain is not merely incremental; it underscores the value of leveraging the graph’s complex relational structure. However, the most critical contribution of this work lies not in the predictive performance itself, but in the validation of the Patient Similarity Network (PSN) as a discovery engine. The topological analysis revealed a profoundly complex structure, evidenced by a Betti number (*β*1) of 63,187. This is not a statistical curiosity; it is mathematical proof that the model learned tens of thousands of non-hierarchical, cyclical patient relationships. The identification of structurally significant features, such as hub nodes (e.g., Patient 817), demonstrates the framework’s ability to autonomously identify potential structural archetypes or computational prototypes. This transforms the PSN from a mere component of a predictive model into a primary object of scientific inquiry, capable of generating concrete, testable hypotheses for further clinical investigation. Furthermore, the discovery of 13 distinct patient clusters validates the model’s capacity to segment the patient manifold into identifiable data-driven sub-regions, providing a discrete map of patient heterogeneity.

The practical value of the framework hinges on its ability to move from an academic exercise to a scalable, enterprise-ready solution. The framework’s architecture was deliberately designed for this purpose, centered on a two-stage process that separates data persistence from computational modeling. The first stage is the architectural foundation: a robust Neo4j graph database serving as the central, persistent repository for complex, interconnected EHR data. The feasibility and performance superiority of this graph-based backend were established in our prior work, which demonstrated up to 48.4x faster query performance over traditional relational databases [45]. The second stage, and the key innovation for bridging research with practice, is the configuration-driven data construction component. This component acts as an intelligent bridge that queries the persistent Neo4j database and dynamically constructs a specialized, in-memory PyTorch Geometric HeteroData object. This object is not just a data container; it is the precise, ML-ready computational graph that the GNN directly consumes for training and inference. This separation of the persistent data store (Neo4j) from the ephemeral computational graph (HeteroData) is the cornerstone of the framework’s modularity and efficiency. It allows feature sets and clinical criteria to be modified via configuration files without altering core model code, which not only facilitates reproducible research but also provides a clear pathway for seamless integration into real-world clinical workflows using interoperability standards like FHIR and SNOMED-CT.

The Patient Similarity Network (PSN) created by this framework serves as more than just a model; it is a structured and computationally efficient knowledge map. This opens a clear pathway toward a next-generation clinical AI that moves beyond simple prediction and towards genuine human-AI collaboration [47–49]. The next step is to leverage this PSN as the grounding knowledge base for a Large Language Model (LLM)-powered clinical co-pilot [53–55]. By using Retrieval-Augmented Generation (RAG), clinicians could interact with this complex data via natural language. However, the ultimate utility of such a system depends critically on the quality of the human-AI dialogue. This is not merely a technical challenge but a cognitive one. The cognitive structure of a prompt—whether it follows a simple, direct reasoning path or a more complex, interactive one— significantly impacts performance and reliability in medical question-answering tasks, a finding established in prior work [46]. The findings suggest that simpler, more direct prompting strategies often yield more stable and trustworthy results in clinical contexts. Therefore, developing a truly effective clinical co-pilot requires a dual approach: grounding the LLM in a factually reliable knowledge base like our PSN, while simultaneously designing the interaction layer based on rigorous, cognitively-informed prompt engineering principles.

Looking further ahead, this human-AI collaborative framework paves the way for a more ambitious future: the autonomous hospital, where intelligent systems are seamlessly integrated into every layer of patient care. In this new era, intelligent physical agents—such as robotic assistants or humanoids—will be integral to diagnostic, surgical, and patient care tasks [50–52]. However, a critical barrier to this vision is safety and reliability; these embodied agents cannot afford the computational latency or the “hallucination” risks of running massive, unconstrained deep learning models in real-time. This is precisely where our work provides a foundational piece. We believe that by grounding their decision-making in a pre-computed and validated knowledge map like our PSN, these agents can make rapid, safe, and contextually aware decisions. This approach moves beyond simple data retrieval; it allows the system to leverage the rich analytical toolkit of graph theory to understand complex patient relationships, identify critical network vulnerabilities, or compute optimal pathways for clinical intervention, all with high computational efficiency [22,35,56,57]. This research thus provides a foundational component for a future where intelligent systems, both digital (as co-pilots) and physical (as assistants), can safely and effectively operate in high-stakes clinical environments, guided by a knowledge system aligned with human cognitive workflows.

This study has several limitations that warrant acknowledgment. First, its retrospective, single-center design means our findings require validation in external, multi-center cohorts to confirm their generalizability. Second, our methodological choices, while practical, offer avenues for future refinement. To maximize data availability, our patient cohort was defined broadly across acute biliary conditions, which may introduce clinical heterogeneity. Future work should therefore focus on more narrowly defined sub-populations, ideally in consultation with clinical specialists Furthermore, it is imperative to recognize that this research was conducted from the perspective of medical informatics and data science, not frontline clinical practice. Consequently, all findings, particularly the interpretations of ‘patient phenotypes’ and ‘clinical archetypes,’ are grounded in computational and statistical patterns rather than direct, patient-facing clinical experience. These data-driven constructs serve as promising starting points for hypothesis generation but lack the essential validation and nuanced contextual understanding that can only be provided by practicing clinicians. Overall, while this study successfully demonstrates the existence of rich topological features, a deep clinical characterization of the identified hubs and cycles was intentionally beyond the scope of this methodological paper. Bridging this gap remains a critical and exciting direction for future interdisciplinary research between data scientists and clinicians.

## 5. Conclusions

This study introduced NETWISE, a framework that successfully recasts GNNs from “black-box” predictors into architects of transparent and interpretable discovery engines. By achieving superior predictive accuracy (AUC 82.5%) and, more importantly, validating the learned patient manifold through both continuous topological analysis and discrete cluster identification, we have established a robust and reproducible methodology for generating data-driven clinical hypotheses. This work provides a foundational methodology for building the reliable, interoperable, and network-based knowledge systems that will underpin the future of intelligent and interactive clinical decision support, ultimately paving the way for safe and effective Human-AI teaming in medicine.

## Data Availability

The MIMIC-IV dataset (version 3.1) used in this study is a publicly available but controlled-access resource The code used for the study is available from the author upon reasonable request.

## Acknowledgements

The author is profoundly grateful to the entire team behind the MIMIC-IV database and the PhysioNet platform. The public availability of this high-fidelity clinical data is an invaluable contribution to the scientific community. This resource removes significant barriers for researchers, particularly those from non-clinical backgrounds. It provides an unparalleled opportunity to gain invaluable, hands-on experience with the intricacies of real-world clinical data and to apply advanced computational methods to solve critical healthcare challenges.

## Author Contribution

The sole author was responsible for all aspects of this work, including conceptualization, methodology, data analysis, and manuscript preparation.

## Data Availability Statement

The MIMIC-IV dataset is accessible via PhysioNet. The code used for the study is available from the author upon reasonable request.

